# Expression predictions inform molecular interpretation of Alzheimer’s disease risk loci

**DOI:** 10.64898/2026.10.07.26364928

**Authors:** ByungHyug Choi, Jaeseung Jeong

## Abstract

Sequence models predict the molecular effects of genetic variation, yet their contribution to interpreting Alzheimer’s disease (AD) association signals remains uncertain. Here we analysed AlphaGenome Atlas predictions for 111,446 common variants across 75 regions from a European-ancestry genome-wide association study (GWAS) of AD and related dementias. Predicted expression effects modestly tracked association within loci (mean partial Spearman ρ = 0.018, *P* = 0.003) and were higher in variants with the highest posterior inclusion probability than in correlated variants outside credible sets (mean difference, 0.044 percentile units; *P* = 0.005). This relationship remained after adjustment for genic position (ρ = 0.011, *P* = 0.026). Predicted splicing effects also tracked association, but this relationship was attenuated after the adjustment. Expression and splicing scores were associated with the corresponding Genotype-Tissue Expression (GTEx) quantitative trait locus (QTL) status, although these analyses did not establish colocalization or agreement in allelic direction. Across 50 loci with multiple fine-mapping candidates, predicted cellular labels were no more concentrated than in position-matched comparisons, and expression direction profiles showed no detectable excess concordance with lead variants. These findings support expression predictions as complementary evidence for interpreting AD risk variants, while showing that the assignment of cellular context must account for fine-mapping uncertainty.

## Introduction

Genome-wide association studies (GWAS) have identified numerous loci contributing to Alzheimer’s disease (AD) and related dementias [1–4]. Translating these discoveries into biological explanations requires identifying the variants that account for association signals, the molecular processes they alter and the cellular contexts in which those alterations matter. Linkage disequilibrium (LD) distributes association across correlated variants, many of which lie outside coding sequence. Statistical fine-mapping reduces uncertainty by assigning each variant a posterior inclusion probability (PIP) and defining credible sets, but several plausible candidates often remain [5]. Biological interpretation must therefore account for uncertainty in variant identity alongside uncertainty in molecular and cellular function.

Functional genomic resources provide complementary evidence for interpreting AD associations. Brain epigenomic maps have implicated regulatory elements active in myeloid cells and microglia, while expression and splicing quantitative trait loci (eQTLs and sQTLs) connect genetic variation to transcript abundance and RNA processing [6–8]. Colocalization evaluates whether disease association and molecular QTL signals may share a causal variant, helping to nominate target genes and mechanisms [4,9]. These findings provide important biological anchors. However, enrichment of a process across disease loci does not establish that the plausible candidates at an individual locus receive a common cellular annotation. When fine-mapping retains several alternatives, the cellular interpretation inferred from one lead variant may depend on which candidate is ultimately causal.

Sequence models offer a systematic way to generate molecular predictions for variants retained by fine-mapping [10–12]. AlphaGenome predicts diverse molecular read-outs, and the AlphaGenome Atlas provides an aggregate variant impact (AVI) score alongside its constituent features [12,13]. Sequence-based annotations have informed complex-trait interpretation [14], while AD applications have examined lead-variant predictions or integrated selected predictions with molecular QTL evidence [15,16]. Such analyses have focused on lead or selected variants, yet alternative candidates within a credible set differ from multiple independent association signals within a locus [17].

Here we evaluate how far sequence predictions support the interpretation of AD association signals, from predicted molecular effects and statistical prioritization to cellular context and expression direction. Using 111,446 common variants in 75 regions from a European-ancestry GWAS of AD and related dementias, we relate predictions for different molecular processes to association strength within loci, compare statistically prioritized candidates with correlated variants outside credible sets, and examine sensitivity to genic position and correspondence with Genotype-Tissue Expression (GTEx) QTL status. We then retain fine-mapping uncertainty when evaluating predicted cellular labels and compare expression direction profiles with those of lead variants. Expression predictions receive the most consistent support across these comparisons, whereas cellular and directional assignments remain unresolved.

## Results

### Study design

We used stage I summary statistics of the European-ancestry AD and related dementias GWAS of Bellenguez et al. [1] (GCST90027158), which included proxy cases defined by family history of dementia. The 75 analysed regions comprised 50 loci identified by genome-wide significant signals in the stage I data and 25 additional regions reported in the source study; within the 25 additional regions, the smallest stage I *P* values among analysed variants ranged from 7.0 × 10^−8^ to 6.3 × 10^−5^ (Methods; Supplementary Table S3). Each region spanned ±250 kb, and together they contained 111,446 biallelic single-nucleotide variants (SNVs) with minor allele frequency ≥1%. APOE and the MHC were excluded a priori because of their atypical association and LD architecture (Methods). For every SNV we obtained AlphaGenome Atlas AVI scores, per-feature AVI attributions and the raw values of the 18 AVI model features, and grouped features into five mechanism classes: expression (RNA-seq, CAGE, PRO-cap, polyadenylation), regulatory (chromatin: DNase, ATAC, histone and transcription-factor ChIP, contact maps), splicing, protein (AlphaMissense and coding-consequence flags) and conservation (Cactus 241-way, PhastCons 470-way). We further obtained track-level RNA-seq and splicing predictions for 371 and 367 cell and tissue tracks.

Before examining the results of each subsequent stage, we documented its hypotheses and analysis decisions in dated protocols. All stages used the same loci and do not constitute independent locus replication. An exploratory stage used AVI attributions. A confirmatory stage fixed the direction of the hypotheses, replaced attributions with raw features, and added fine-mapping, genic-position and GTEx analyses. A cellular-context stage examined whether the credible-set candidates of a locus receive a common predicted cellular label and whether their predicted direction profiles agree with that of the lead variant (Methods).

### Aggregate variant impact tracks association on average but not uniformly

Within each locus we computed the partial Spearman correlation between AVI and association χ^2^, adjusting for LD score, minor allele frequency and distance to the nearest transcription start site (TSS), and compared the mean over loci with a null that circularly shifts the annotation within each locus. AVI increased with association strength (mean partial correlation ρ = +0.013; null 95th percentile +0.005; one-sided *P* = 0.001; Fig. 1a). The correlation was positive in 64% of loci (49% under the null), but loci differed more than expected by chance (heterogeneity *P* = 0.002): some loci showed strong concordance (for example L019, L038 and L039 at chr6:47.63, chr7:54.88 and chr19:54.27 Mb; *z* > 2.9) and others the opposite (*z* < −2.2).

**Figure 1.**
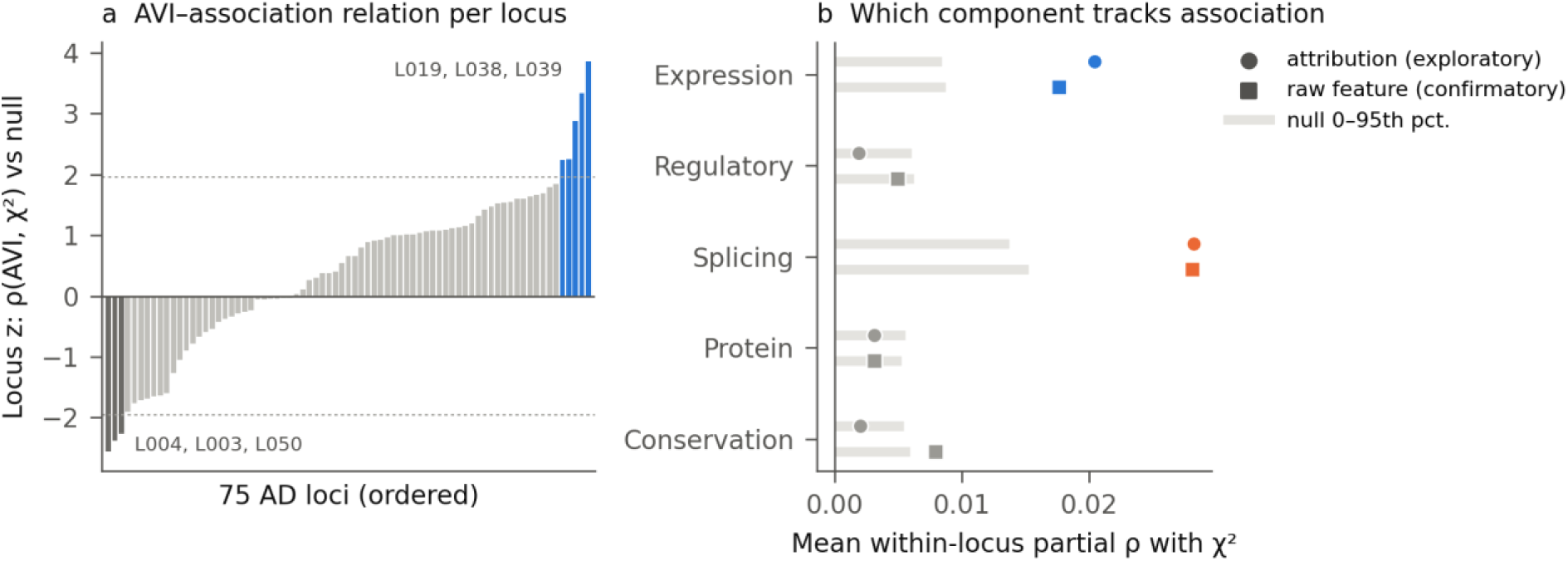
Aggregate variant impact and its components versus association within AD loci. **a**, Per-locus *z* score of the partial Spearman correlation between AVI and association χ^2^ relative to a within-locus circular-shift null (75 loci; identifiers as in Supplementary Table S3). Dotted lines, |*z*| = 1.96. Mean ρ = +0.013 (*P* = 0.001); heterogeneity *P* = 0.002. **b**, Mean within-locus partial correlation for each mechanism-class score, using AVI attributions (circles, exploratory stage) and raw AVI features (squares, confirmatory stage). Grey bars span 0 to the 95th percentile of the circular-shift null. Expression (blue) and splicing (orange) exceed the null in both stages.

### Association is concentrated in the expression and splicing components

AVI attributions were dominated by conservation (mean share 0.47) and regulatory features (0.34), with smaller contributions from expression (0.12) and splicing (0.07). Yet the association was tracked by the minor components: expression (ρ = +0.020, *P* = 0.001) and splicing (ρ = +0.028, *P* = 0.004), but not regulatory (+0.002, *P* = 0.29), protein (+0.003, *P* = 0.17) or conservation (+0.002, *P* = 0.25) (Fig. 1b, circles).

Attributions proved to be a poor mechanism label: a positive control, the common *PILRA* missense variant G78R (rs1859788) [18], had a high AVI (99.4th percentile) but only 3% of its attribution in the protein class. We therefore re-measured each class using the raw AVI model features. Raw-feature class scores passed pre-specified validity checks (Supplementary Fig. S1): the protein score separated variants annotated as missense by the Ensembl Variant Effect Predictor (VEP) from other variants (AUC 0.88; 1,260 missense variants), the splicing score separated splice-region variants from other genic variants (AUC 0.78; 767 splice-region variants), and G78R ranked in the 99.1st percentile for the protein score. With raw features and pre-specified one-sided hypotheses, both expression (ρ = +0.018, *P* = 0.003) and splicing (ρ = +0.028, *P* = 0.002) again tracked association (Fig. 1b, squares). Conservation showed a weaker relationship (+0.008, *P* = 0.015; not pre-specified).

The relationship was concentrated at the top of the association distribution (Fig. 2). Across within-locus χ^2^ deciles, expression and splicing scores were flat through the eighth decile and rose in the ninth and tenth, with the largest rise for splicing (mean percentile 0.508 in decile 1 vs 0.550 in decile 10). These small average correlations indicate a shift in predicted scores among more strongly associated variants; they do not imply that highly associated variants within a locus share one dominant molecular mechanism.

**Figure 2.**
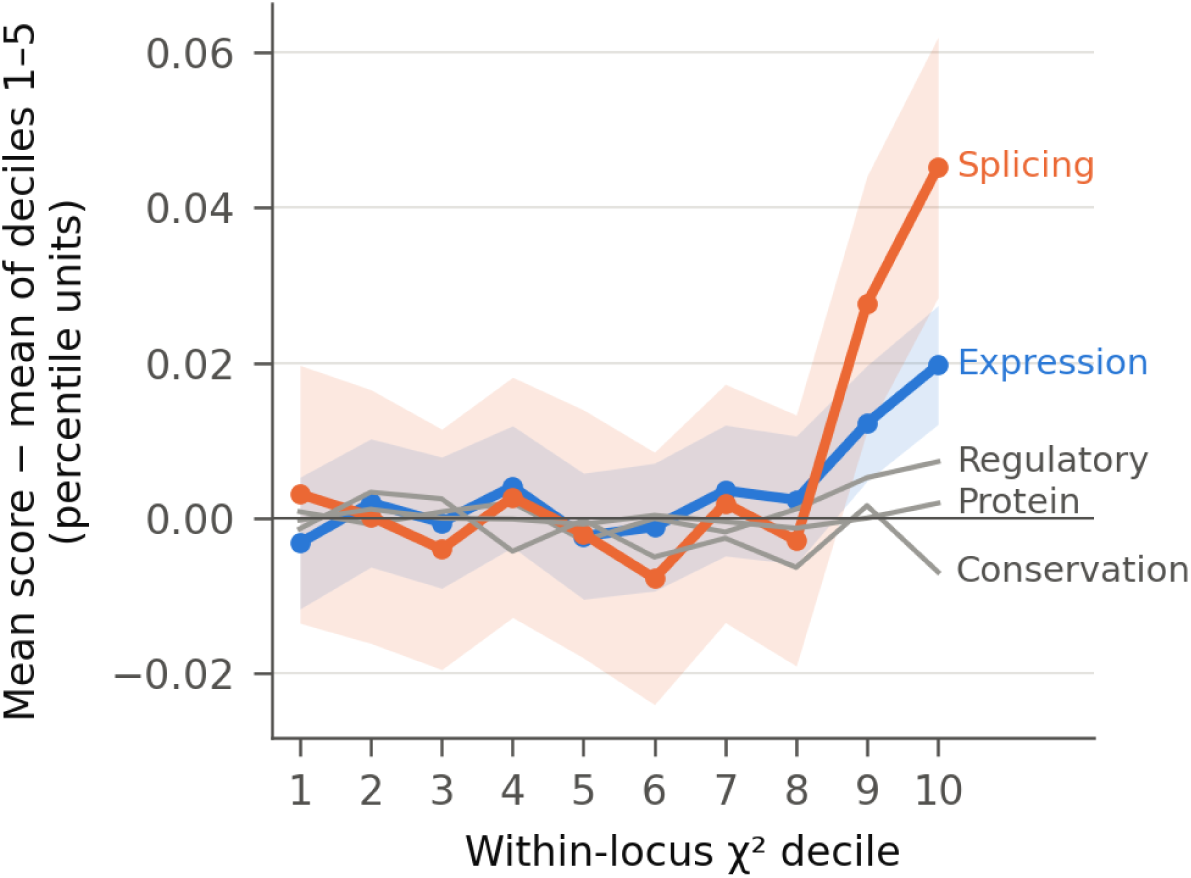
Class scores across within-locus association deciles. Mean raw-feature class score (percentile units) per within-locus χ^2^ decile, averaged over 75 loci and centred on the mean of deciles 1–5. Shading, ±1 s.e.m. across loci for expression and splicing.

### Highest-PIP variants have higher predicted expression scores than correlated variants outside credible sets

To ask whether the relationship reflects the variants prioritized by fine-mapping rather than their correlated neighbours, we fine-mapped each locus with SuSiE-RSS using a uniform prior, so that fine-mapping used no AlphaGenome information [5,19]. This yielded 95 credible sets in 63 loci (median size 6). For each credible set, we compared the highest-PIP variant with correlated variants outside any credible set (*r*^2^ ≥ 0.5 with the highest-PIP variant); 87 credible sets had at least one such variant. Highest-PIP variants had higher expression scores than correlated variants outside any credible set (mean difference, 0.044 percentile units; 67% of 87 comparisons positive; sign-flip *P* = 0.005; Fig. 3a). Splicing scores did not differ (mean difference, 0.003; *P* = 0.43). Regulatory scores were also higher in highest-PIP variants (mean difference, 0.046; *P* = 0.033; not pre-specified), suggesting that part of this difference reflects general predicted regulatory activity.

**Figure 3.**
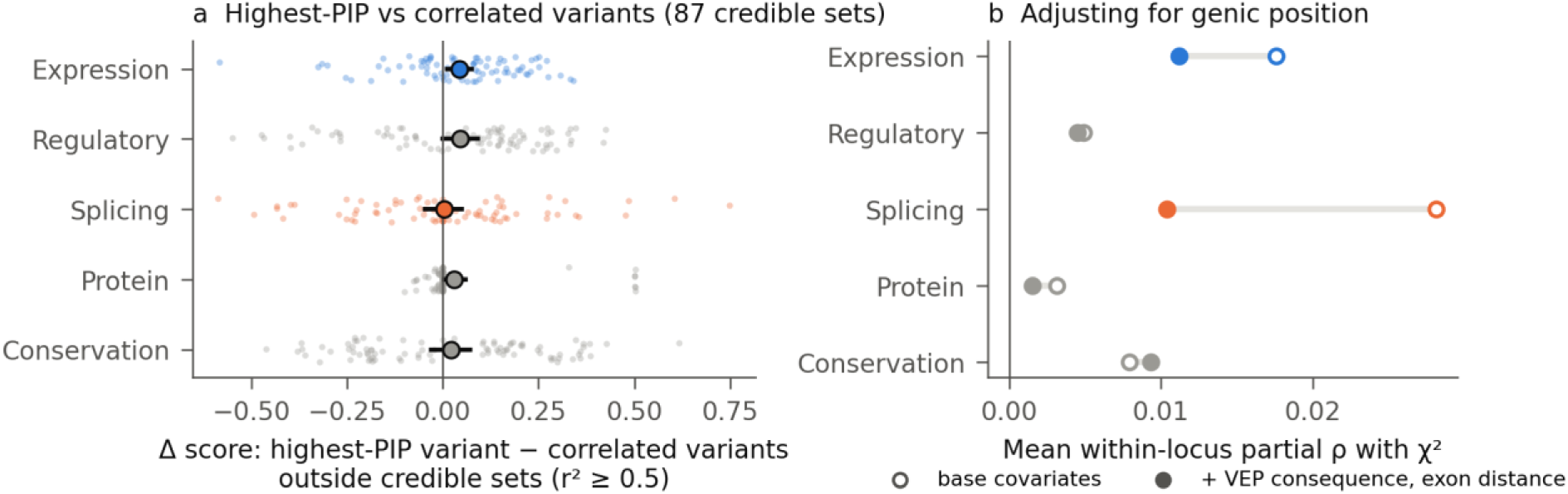
Highest-PIP variants versus correlated variants outside credible sets, and control for genic position. **a**, For each of 87 uniform-prior SuSiE credible sets, difference between the class score of the highest-PIP variant and the mean score of correlated variants outside any credible set (*r*^2^ ≥ 0.5). Small points, credible sets; large points, mean with bootstrap 95% CI. **b**, Mean within-locus partial correlation with χ^2^ before (open) and after (filled) additional adjustment for VEP consequence category and distance to the nearest exon boundary.

### The expression signal is not explained by genic position

Associated variants could have higher expression or splicing scores simply because they lie in or near genes. We added VEP consequence categories (coding, UTR, splice region, intronic, flanking, non-coding exon, intergenic) and distance to the nearest exon boundary to the covariates. The expression relationship was attenuated but persisted (ρ = +0.011, one-sided *P* = 0.026), whereas the splicing relationship was reduced by about two-thirds and was no longer significant (+0.010, *P* = 0.14; Fig. 3b). When the top 1% of variants in each locus were compared with background variants re-weighted to the same consequence composition, the top set remained enriched for expression (*P* = 0.026), splicing (*P* = 0.036), regulatory (*P* = 0.008) and protein (*P* = 0.036) scores (two-sided tests).

### Predicted expression and splicing scores are associated with corresponding GTEx QTL status

We queried GTEx v8 [7] for all 175 credible-set variants with PIP ≥ 0.1; 83% had significant eQTL status and 51% had significant sQTL status in at least one tissue. In joint logistic models adjusting for minor allele frequency and TSS distance, the expression score was associated with eQTL status (odds ratio [OR] 1.73 per SD, 95% CI 1.07–2.80; one-sided *P* = 0.013) and the splicing score with sQTL status (OR 1.50, 1.04–2.16; *P* = 0.014; Fig. 4). Neither cross term showed a positive association: the expression score was not associated with sQTL status (OR 1.01), and the splicing score was, if anything, inversely related to eQTL status (OR 0.56). These modality-consistent associations do not establish that the AD and QTL signals share a causal variant, nor do they test whether predicted and observed allelic effects have the same direction.

**Figure 4.**
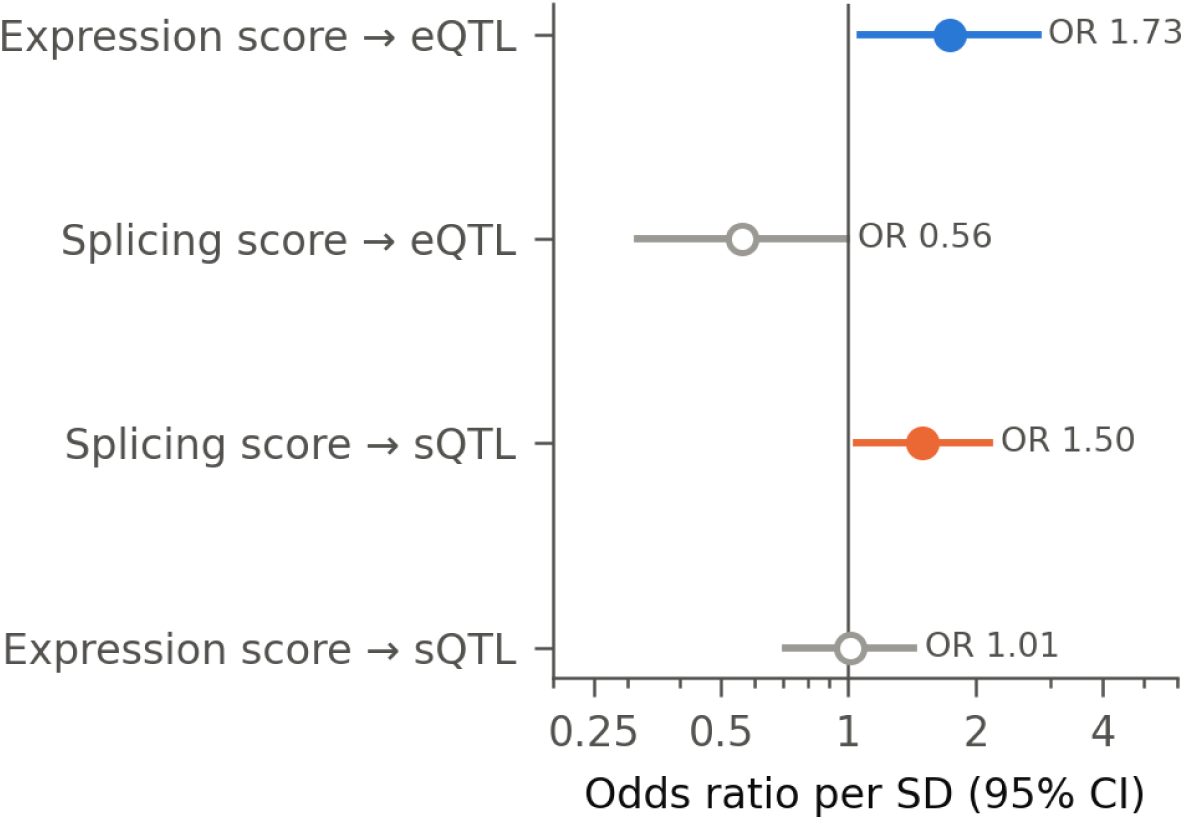
AlphaGenome class scores and GTEx molecular QTL status of credible-set variants. Odds ratios per SD from joint logistic models (both class scores, minor allele frequency and log TSS distance) for being a significant GTEx v8 eQTL or sQTL in any tissue (*n* = 175 variants, PIP ≥ 0.1). Filled, matched class and QTL type; open, cross terms. The analysis concerns QTL status only; it does not test colocalization or the direction of allelic effects.

### Top associated variants shift towards expression and splicing without sharing a dominant mechanism

We next compared the top 1% of variants in each locus with the rest of the locus. The top sets shifted in a consistent direction relative to their loci, towards expression (*P* = 0.016), splicing (*P* = 0.036) and protein (*P* = 0.018) and away from conservation (*P* = 0.032), and the direction of this shift was more similar across loci than expected (mean cosine +0.037 vs +0.001; *P* = 0.018). In the exploratory attribution-based analysis, dominant mechanism labels did not show increased agreement among the top associated variants within individual loci (0.46 versus 0.50 in position-matched sets), including among variants in low LD. At the level of individual loci, seven showed a nominal shift (*z* > 1.64) towards splicing or expression in raw-feature class scores after re-weighting for consequence composition (Table 1). These include *GRN*, whose highest-PIP variant rs5848 (PIP 1.00) lies in the 3`′` UTR [20], and *TSPAN14*, whose credible-set variants are GTEx sQTLs for *TSPAN14*, a TspanC8 tetraspanin that regulates ADAM10 [21]. Single-locus classifications have low power and are intended as hypotheses.

**Table 1.** Loci with a nominal shift of top-associated variants towards expression or splicing. *z*, shift of the top-1% set in raw-feature class scores relative to background variants re-weighted to the same consequence composition, standardized against a position-preserving null. Top-PIP variant, highest-PIP variant of the uniform-prior credible set (PIP in parentheses); –, none. GTEx genes for credible-set variants with PIP ≥ 0.1.

| Locus (hg38) | Nearest genes | $z_{\text{expr}}$ | $z_{\text{splice}}$ | Top-PIP variant (PIP) | GTEx eQTL | GTEx sQTL |
| --- | --- | --- | --- | --- | --- | --- |
| chr17:44.35 Mb | *GRN*, *FAM171A2* | -0.3 | 2.3 | rs5848 (1.00) | *GRN*, *FAM171A2*, *ITGA2B* | *ITGA2B* |
| chr10:80.49 Mb | *TSPAN14* | 1.1 | 2.1 | chr10:80494228 (0.12) | *TSPAN14*, *ANXA11*, *DYDC2*,<br>*FAM213A*, *LINC00857*,<br>*MAT1A*, *PLAC9*,<br>*RP11-137H2.4*, *SFTPD* | *TSPAN14*,<br>*DYDC1* |
| chr6:47.63 Mb | *CD2AP*, *ADGRF4* | 1.1 | 1.7 | chr6:47627419 (0.07) | – | – |
| chr3:155.08 Mb | *MME* | 1.2 | 1.6 | chr3:155084189 (0.60) | – | *SLC33A1* |
| chr8:100.66 Mb | *SNX31*, *PABPC1* | 1.7 | 0.2 | chr8:100663356 (0.26) | *PABPC1*, *SNX31* | – |
| chr4:40.20 Mb | *RHOH* | 2.0 | 0.3 | chr4:40197226 (0.99) | *RHOH*, *RP11-395I6.3* | – |
| chr2:105.75 Mb | *NCK2* | 1.9 | 1.6 | – | – | – |

### Predicted cellular labels of credible-set candidates depend on which candidate is assumed to be causal

Because expression and splicing scores tracked association, we next asked whether the credible-set candidates of a locus receive a common predicted cellular label. For each SNV we took the Atlas RNA-seq predictions across 371 tracks (285 cell types and tissues), expressed as genome-calibrated quantiles, and grouped the tracks into 12 pre-specified cell classes (Methods). The predicted cellular label of a variant was the class with the largest predicted effect after calibrating each class against its own distribution across all 111,446 SNVs. This calibration reduces the advantage of classes with many tracks: before calibration 31% of variants were assigned to the epithelial/other class, and after calibration the class shares ranged from 4.7% to 12.7%. For each locus we summarized the PIP-weighted distribution of predicted cellular labels among credible-set variants with the Shannon entropy, where lower entropy indicates greater concentration on fewer labels.

Across 50 loci, predicted cellular labels of credible-set candidates were not more concentrated than those of variants matched by position (mean PIP-weighted entropy, 1.63 versus 1.53; one-sided *P* for greater convergence = 1.00; Fig. 5a,b). The comparison sets were the credible sets moved to other positions in the same locus with their spacing preserved. Thirty-nine of the 50 loci lay above the diagonal in Fig. 5a, and no locus showed nominally greater concentration (2.5 expected under the null); the closest was chr5:180.2 Mb (78% blood/immune; *P* = 0.10). Predicted labels of candidates within the same locus spanned brain, immune, hepatic and metabolic, vascular and other classes (Fig. 5c). The result was the same when cell class and mechanism (expression or splicing) were combined into 24 categories (entropy 1.78 vs 1.64; *P* = 1.00). It was also the same when each variant contributed its top three classes, when the calibration was omitted, and when only variants with PIP ≥ 0.1 were used (*P* = 0.66–1.00).

**Figure 5.**
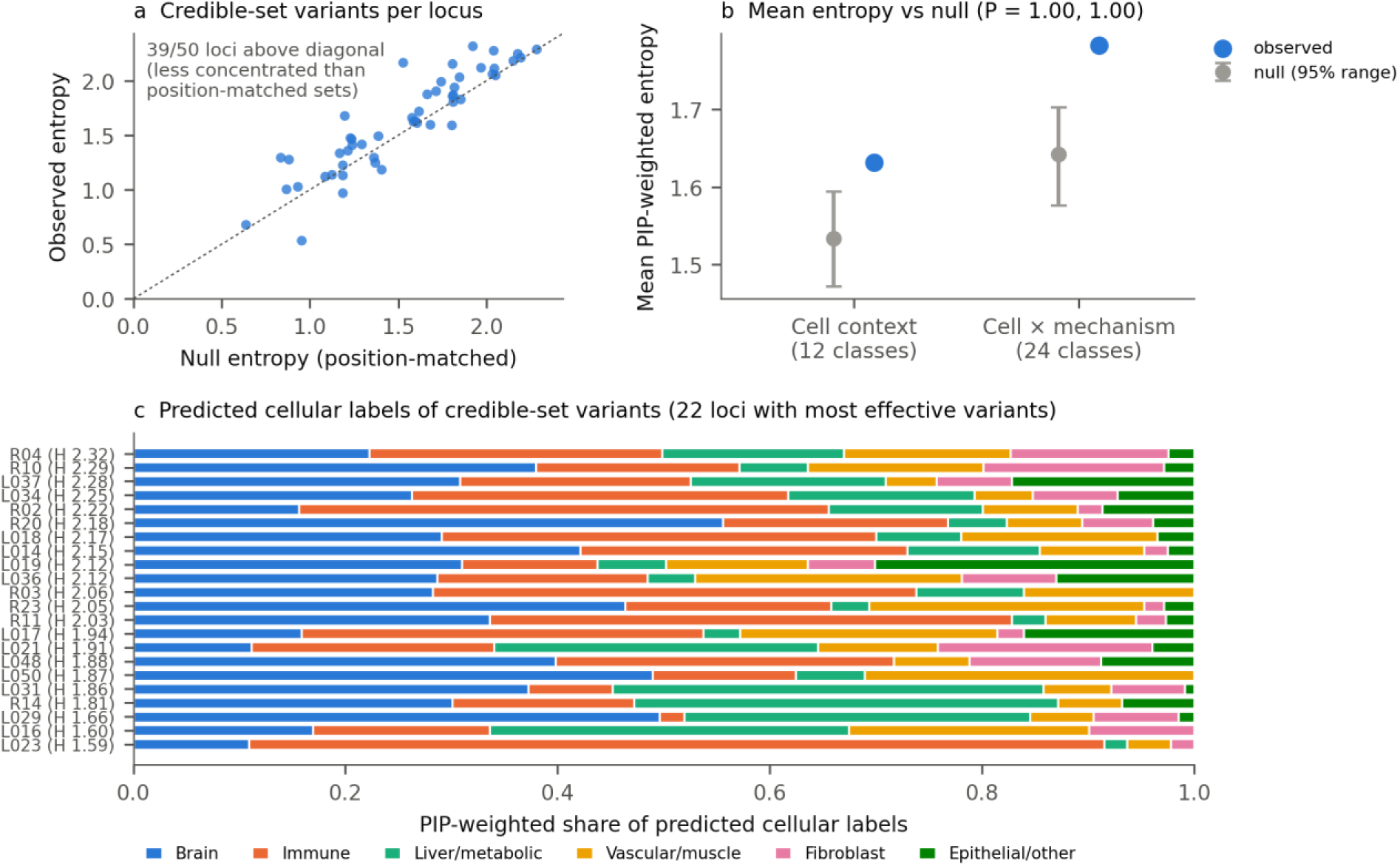
Credible-set candidates do not resolve a common predicted cellular context. **a**, PIP-weighted entropy of the predicted cellular labels (12 classes) of credible-set variants in each of 50 loci versus the mean entropy of position-matched sets. Points above the diagonal (39 of 50) are less concentrated than the position-matched sets. **b**, Mean entropy over loci for cellular labels (12 classes) and cell × mechanism labels (24 classes). Grey, null mean and 95% range (1,000 replicates). One-sided *P* for greater concentration = 1.00 for both. **c**, PIP-weighted share of predicted cellular labels among credible-set variants for the 22 loci with the largest effective number of variants, ordered by entropy *H*; locus identifiers as in Supplementary Table S3. The 12 classes are grouped into six super-classes for display.

Secondary measures, including measures that do not depend on the class definitions, gave the same answer. Pairs of top-1% associated variants agreed on the class of their most affected track less often than position-matched random sets (0.20 vs 0.22; *P* = 0.97). The same was true for splicing predictions (0.29 vs 0.36; *P* = 1.00), for the exact biosample of the most affected track (0.024 vs 0.055; *P* = 1.00) and for the rank order of all 371 tracks (mean pairwise Spearman ρ, 0.13 vs 0.16; *P* = 1.00). Among pairs in low LD (*r*^2^ < 0.1), agreement did not exceed that of position-matched sets for class (0.16 vs 0.19; *P* = 0.86) or track rank order (0.12 vs 0.11; *P* = 0.36; Supplementary Table S4). Representative variants of different credible sets in the same locus agreed more often than variants from different loci (0.19 vs 0.09; *P* = 0.026). They did not agree more often than random SNVs from the same locus (0.15; *P* = 0.22, post hoc). This pattern is consistent with a shared locus context, such as the same nearby genes, rather than with greater concentration of predicted labels.

### Predicted expression direction profiles of credible-set candidates show no detectable concordance with the lead

Candidates could receive their largest predicted effects in different tracks yet still share a signed pattern of predicted increases and decreases across tracks. We therefore compared signed predictions. We selected the GWAS lead variant of each locus (minimum stage I *P*), credible-set members, the top 1% of associated variants and random variants (9,267 SNVs in total). We oriented the Atlas signed quantiles to the AD risk allele. For the gene most affected by the lead variant, we then correlated each variant’s 371-track risk-oriented profile with that of the lead (ρ_dir_).

Signed AlphaGenome predictions varied across RNA-seq tracks for many variants. For each SNV’s most affected gene, 54% of SNVs had the minority sign in at least 10% of strongly affected tracks (|quantile| ≥ 0.5), and only 10% had the same sign in all such tracks (Fig. 6a). The mean profile correlation for credible-set members was 0.001, compared with 0.006 for variants matched by distance, providing no evidence of greater concordance with the lead (one-sided *P* = 0.62; Fig. 6b). The top 1% of variants outside credible sets were likewise no more concordant with the lead than matched variants (+0.003 vs +0.001; *P* = 0.45). Variants in high LD with the lead (*r*^2^ ≥ 0.5) were no more concordant with the lead than matched variants (−0.004 vs +0.008; *P* = 0.75), and variants in moderate or low LD showed a weak, non-significant positive tendency (+0.033 vs +0.001; *P* = 0.13–0.14). Results were unchanged with sign concordance as the metric (0.501 vs 0.501) and with genes strongly affected by both variants.

**Figure 6.**
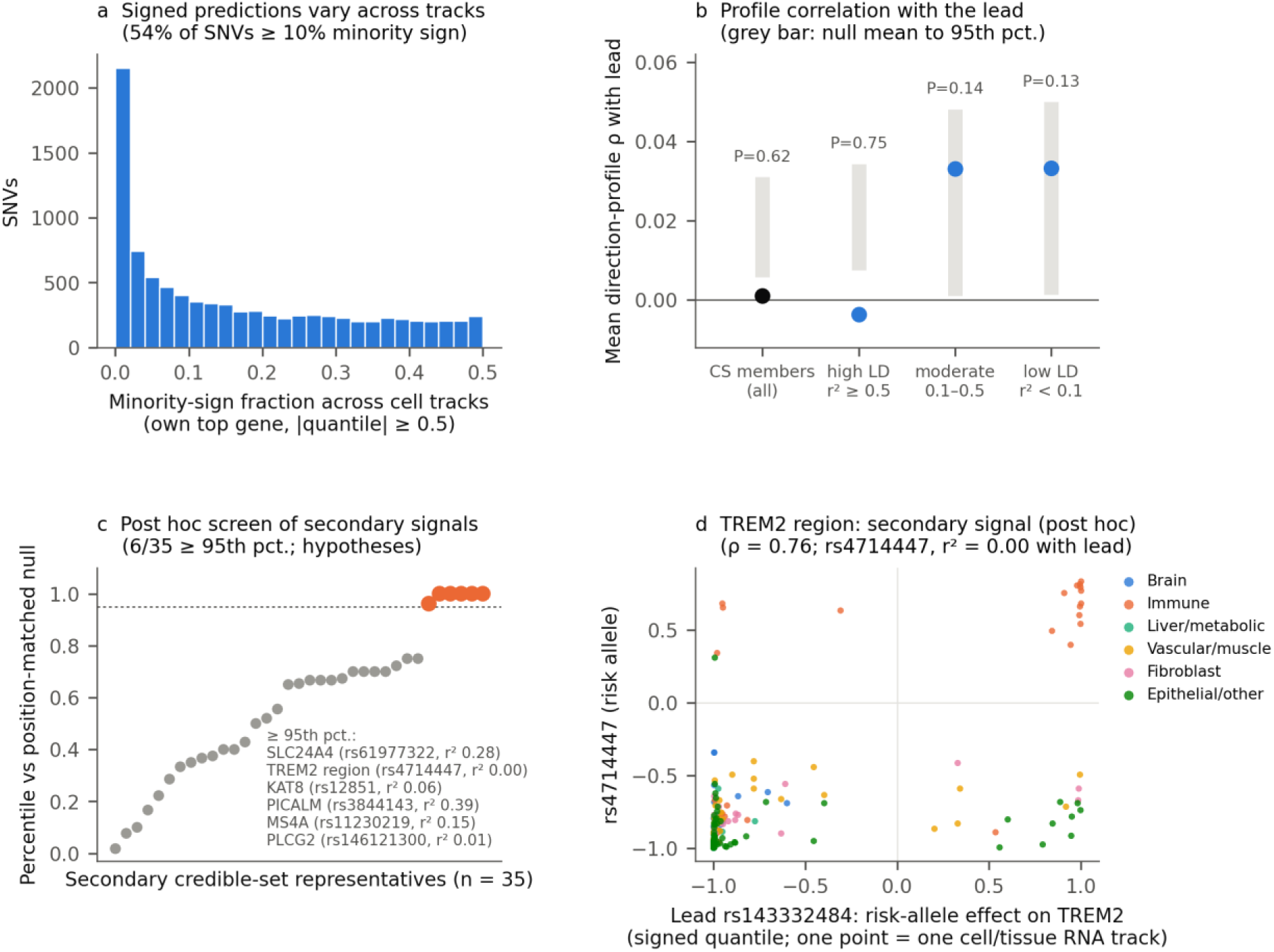
Predicted expression direction profiles across RNA-seq tracks. **a**, For each of 9,247 SNVs with at least five strongly affected tracks (|quantile| ≥ 0.5) for its most affected gene, the fraction of those tracks with the minority sign. **b**, Mean Spearman correlation between the 371-track, risk-allele-oriented profiles of the lead and of credible-set members, overall and by LD with the lead. Grey bars span the mean to the 95th percentile of the distance-matched null; one-sided *P* values. **c**, Percentile of ρ_dir_ for the representative variant of each of 35 secondary credible sets relative to distance-matched random variants of the same locus (median 12 per representative). Orange, ≥95th percentile in a post hoc screen (6 observed, 3.3 expected by chance; exact *P* = 0.11); these loci are hypotheses for validation. **d**, *TREM2* region: risk-allele-oriented signed quantiles of the predicted effect on *TREM2* for the lead rs143332484 (x axis) and the secondary signal rs4714447 (y axis). Each point is one of 371 RNA-seq tracks, coloured by cell super-class. Positive values indicate predicted up-regulation by the risk allele, and values near ±1 indicate predictions in the genome-wide extreme; these are model predictions, not measured effects.

In a post hoc screen, we compared the representative (highest-PIP) variant of each of 35 secondary credible sets (those not containing the lead) with the lead. Six exceeded the 95th percentile of their distance-matched comparison distributions, in the *TREM2* region and at the *SLC24A4*, *PICALM*, *MS4A*, *PLCG2* and *KAT8* loci (Fig. 6c and Supplementary Table S2). Because each percentile was computed against few matched variants (median 12), 3.3 were expected by chance, and the excess was not significant (exact *P* = 0.11). Two of the six (*MS4A* and *KAT8*) lie in loci with suspected LD mismatch, where 10 of 11 secondary credible sets consisted of a single variant with PIP ≈ 1. The clearest case was the *TREM2* region: rs4714447, which is not in LD with the lead rs143332484 (*r*^2^ < 0.01), had a predicted *TREM2* profile that correlated with that of the lead more strongly than all 56 matched variants (ρ**_dir_** = 0.76; Fig. 6d); for both variants, the risk allele was predicted to increase *TREM2* expression in immune tracks and to decrease it in most other tissues. These are model predictions and do not show that either variant regulates *TREM2* in the same direction in cells. Moreover, rs143332484 is the *TREM2* missense variant p.Arg62His (R62H) [22], whose established effect is on protein function. In five of the six loci, the lead’s most affected gene was not the gene after which the locus is named (for example, *CHGA* at *SLC24A4* and *RNU6-560P* at *PICALM*; Supplementary Table S2).

### Exploratory comparison with microglial MPRA

Finally, we asked whether the credible-set member with the highest expression score was more often identified as an allelic regulatory variant in a microglial massively parallel reporter assay (MPRA) [23] than the GWAS lead. This comparison was underpowered (25 credible sets, five discordant pairs) and inconclusive. Across all tested variants, expression scores were associated with allelic activity in HMC3 cells (AUC 0.64, *P* = 0.001) but not in iPSC-derived microglia, the primary model (AUC 0.48; Supplementary Note 1).

## Discussion

Association strength showed modest relationships with predicted transcript effects that were supported by several analyses, most clearly for expression, whereas the predicted cellular labels and direction profiles of fine-mapping candidates differed within loci. Predicted chromatin and protein-coding components did not track association, and conservation showed at most a weak relationship that was not pre-specified.

The expression relationship was supported by four complementary analyses. It tracked association within loci, it was stronger in highest-PIP variants than in correlated variants outside credible sets, it persisted after adjustment for genic position, and the expression score was associated with GTEx eQTL status. This pattern resembles multi-ancestry benchmarks in which sequence-model predictions agreed weakly with nominal eQTL effect sizes but better discriminated high-PIP fine-mapped variants [24]. The splicing relationship should be interpreted separately. Splicing scores tracked association and were associated with sQTL status, but they did not differ between highest-PIP variants and correlated variants outside credible sets, and the within-locus relationship was reduced by about two-thirds and was no longer significant after adjustment for genic position (ρ = +0.010, *P* = 0.14). We therefore interpret the splicing signal as largely regional, although the top 1% of variants remained enriched for splicing scores after re-weighting for consequence composition: association at AD loci concentrates in genic regions, where predicted splicing effects are large. This does not exclude splicing mechanisms, which are a major link between genetic variation and disease [8], but our data do not distinguish them from genic position. None of these analyses tested whether predicted and observed allelic effects have the same direction.

Because variants within a credible set are alternative explanations for an association signal, diversity in their predicted cellular labels does not demonstrate that multiple causal variants act in different cell types. It shows instead that, with current predictions, the cellular label of a locus depends on which candidate is assumed to be causal, so a cellular annotation inferred from a lead variant may not describe the locus once fine-mapping uncertainty is taken into account. Aggregating predicted effects of lead variants across loci has shown enrichment in peripheral immune and metabolic programmes [15], and epigenomic studies have implicated microglial enhancers [6]; such enrichment across loci is compatible with our results, which concern the assignment of labels to candidates within a locus. The entropy analysis pooled the candidates of all credible sets in a locus, so label diversity can reflect alternative candidates within one credible set or distinct independent signals; these are different situations [17], and our analysis does not separate them. Label diversity may also reflect differences between candidates in the gene with the largest predicted effect, the presence of several genes within AD loci, or noise in variant-level predictions.

Direction profiles showed no detectable concordance between credible-set members and the lead, including for variants in high LD with the lead, which are statistically interchangeable with it; as with cellular labels, the predicted direction of effect therefore depends on which candidate is assumed to be causal. In a post hoc screen, secondary signals with concordant profiles were no more frequent than expected by chance, but the *TREM2* region stood out (ρ**_dir_** = 0.76 with a variant not in LD with the lead); this locus is a hypothesis for subsequent validation rather than evidence that the two signals regulate the same gene in the same direction. Weak predicted effects can yield inconsistent signs across tracks and between variants without reflecting opposing biological effects [25], so the absence of direction concordance does not show that candidates act in opposite directions. At *TREM2*, the lead is a coding variant, so any shared expression effect would add to, not replace, its protein-level effect.

The finding that the two components dominating the aggregate score, conservation and chromatin, tracked association weakly or not at all is methodologically relevant. This does not show that chromatin mechanisms are uninvolved. Microglia, in which AD risk variants are enriched in enhancers [6], are not represented in the Atlas tracks, and the regulatory class pools many assays and cell types, which can dilute a cell-type-specific signal. Prediction accuracy also differs between modalities [26], so differences between mechanism classes reflect model performance and score construction as well as biology. Aggregate variant-impact scores and their attributions are optimized to rank variant deleteriousness broadly [13], and in our data attributions obscured even a known missense mechanism. Mechanism-specific raw features were better suited to linking GWAS signals to molecular processes. The heterogeneity across loci argues against applying a single functional prior uniformly, and a lead variant’s cellular annotation should not be assigned to an entire locus without accounting for fine-mapping uncertainty.

Several limitations apply. First, all stages used the same 75 loci and do not constitute independent locus replication. The confirmatory stage fixed hypothesis directions, changed the measurement, and added GTEx data and fine-mapping that did not use AlphaGenome information. Testing the same hypotheses in independent, sub-threshold loci remains necessary. Second, effect sizes are small (ρ ≈ 0.02), as expected when most variants in a locus are non-causal; they describe a shift in the distribution of association, not a variant-level predictor. Sequence models may also underestimate the predicted effects of causal eQTL variants relative to nearby variants [25], which would attenuate these correlations. Third, fine-mapping used a 1000 Genomes European reference panel of 525 individuals, and LD mismatch with the GWAS sample may affect credible sets (excluding two loci with evidence of mismatch did not change the within-locus results). Fourth, neither the AlphaGenome Atlas nor GTEx includes microglia, and monocytes and other myeloid tracks are proxies. Single-nucleus eQTL maps of the aged human cortex, which include microglia [27], provide a cell-type-resolved benchmark that is not represented in GTEx or the current Atlas tracks. The exploratory MPRA comparison was inconclusive in iPSC-derived microglia, despite an association between expression scores and allelic activity in HMC3 cells; therefore, it does not provide consistent microglial validation of variant prioritization (Supplementary Note 1). Fifth, the GTEx analyses tested QTL status only; they did not test GWAS–QTL colocalization or whether predicted and observed allelic effects have the same direction. Sixth, the cell-class mapping involves judgement; class-free measures (exact biosample and full track rank order) likewise showed no greater agreement than position-matched sets (Supplementary Table S4). The predicted cellular label was defined by the class with the largest predicted effect over all genes, so it reflects both the cell type and the gene most affected. Direction profiles were computed for a single target gene, which at several loci was a non-coding or unannotated gene. Finally, the Bellenguez stage I GWAS is of European ancestry and includes AD-related dementias and proxy cases, which may dilute AD-specific signals; moreover, agreement between sequence-model predictions and observed eQTL effects varies with ancestry and fine-mapping resolution [24].

In summary, in the AD regions analysed here, expression predictions received the most consistent molecular support, whereas cellular and directional assignments remained uncertain once plausible candidates were retained. Sequence-based variant prioritization should therefore operate on fine-mapped candidates rather than on lead variants alone. Reporting predicted molecular effects together with uncertainty in candidate identity and cellular context provides a clearer basis for selecting hypotheses for functional evaluation.

## Methods

### GWAS data and loci

We used stage I summary statistics of Bellenguez et al. [1] (GWAS Catalog GCST90027158; GRCh38), a European-ancestry GWAS of AD and related dementias that included proxy cases. Primary loci were defined by iterative distance-based clumping of variants with *P* < 5 × 10^−8^: around each successive minimum-*P* variant we removed ±1 Mb and took ±250 kb as the locus. This gave 50 loci after two regions were excluded. The APOE locus (chr19:43.9–45.9 Mb) was excluded a priori because of its exceptionally large association effect and established coding-driven risk architecture, which could disproportionately influence analyses intended to characterize functional variation across typical AD loci. The MHC region (chr6:25–34 Mb) was excluded because of its unusually complex and extended LD structure. We added 25 regions from associations reported for the same study in the GWAS Catalog that lay more than 1 Mb from the 50 leads (signals within 1 Mb merged). Within these 25 regions, the smallest stage I *P* values among analysed variants ranged from 7.0 × 10^−8^ to 6.3 × 10^−5^; the Catalog-reported leads of six regions were not analysed because three had a minor allele frequency below 1% and three were insertions or deletions (Supplementary Table S3). Within each window we kept biallelic SNVs with minor allele frequency ≥1%, sample size ≥80% of the maximum, and presence in the reference panel. Association χ^2^ = (β/SE)^2^.

### Reference panel and covariates

LD was computed from unrelated European individuals (*n* = 525) of the high-coverage 1000 Genomes Project on GRCh38 [28]. LD score was the sum of *r*^2^ with all SNVs within ±250 kb. Gene, exon and variant-consequence annotations were obtained through the Ensembl REST API (release 116; accessed 24–26 September 2026). TSS distance was to the nearest Ensembl protein-coding TSS. Variant consequences were obtained with the Ensembl VEP REST service [29] (most severe consequence; missense and splice-site flags), and exon-boundary distance from Ensembl exon coordinates.

### AlphaGenome Atlas features and class scores

We queried the AlphaGenome Atlas [12,13] through the alphagenome Python client (version 0.9.0) for the scorers AVI_SCORE, AVI_SCORE_FEATURE_IMPORTANCE (per-feature attributions) and AVI_SCORE_MODEL_FEATURES (raw feature values). The 18 features were grouped into classes as listed in Results; insertion/deletion flags are zero for SNVs and were not used. In the exploratory stage, a class score was the sum of absolute attributions of its features. In the confirmatory stage, each raw feature was converted to a percentile across all 111,446 SNVs, and the class score was the maximum percentile over its features. Percentiles used absolute values for modality maxima, splicing and AlphaMissense [30], with missing values set to zero; raw values for conservation [31,32]; and 0/1 for coding-consequence flags.

### Within-locus association with annotations

For each locus we computed the partial Spearman correlation between a score and χ^2^, adjusting for rank-transformed LD score, minor allele frequency and log TSS distance, and averaged over loci (ρ). The null distribution was obtained by circularly shifting the score along the ordered SNVs within each locus by a random offset (1,000 replicates; the same replicate index across loci), which preserves the local correlation structure of the annotation. Heterogeneity was assessed with *H* = Σ(*z*_ℓ_ − *z*)^2^, where *z*_ℓ_ standardizes each locus correlation against its own null, and compared with the same statistic in null replicates (two-sided). For the genic-position analysis, dummy-coded VEP consequence categories and log distance to the nearest exon boundary were added to the covariates. Sensitivity analyses excluded two loci with suspected LD mismatch, at which SuSiE-RSS did not converge (L006 and L028; Supplementary Table S3), and used the AVI quantile instead of AVI.

### Top-set mechanism classes

For each locus, the top set was the 1% of SNVs with the largest χ^2^ (at least five). Dominant-class agreement was the fraction of top-set pairs with the same argmax attribution class; class shift was the difference between top-set and locus-wide mean class shares. The null translated the whole top set by a random offset within the locus (circularly) and replaced each position by the nearest SNV, preserving inter-variant distances (1,000 replicates). Pairs were stratified by LD (*r*^2^ ≥ 0.5 vs < 0.1). Cross-locus consistency was the mean pairwise cosine of locus shift vectors. For the genic-position analysis, the raw-feature class scores of the top set were compared with those of background variants re-weighted to the consequence composition of the top set (two-sided tests). Per-locus *z* scores (Table 1) standardized this shift for each locus and class against its own 1,000 translations of the top set.

### Fine-mapping and comparison with correlated variants

We ran SuSiE-RSS [5,19] (susieR 0.12.35) with *L* = 10, coverage 0.95, minimum absolute correlation 0.5, the median sample size of the locus, fixed residual variance and a uniform prior. For each credible set, we compared the highest-PIP variant with correlated variants, defined as SNVs outside any credible set with *r*^2^ ≥ 0.5 to the highest-PIP variant. The statistic was the mean over credible sets of (highest-PIP variant score − mean score of its correlated variants), with a null from random sign flips of the per-set differences (10,000 permutations). Bootstrap intervals in Fig. 3a resample credible sets.

### GTEx analysis

For credible-set variants with PIP ≥ 0.1 we retrieved significant single-tissue eQTL and sQTL associations from the GTEx v8 portal API [7]. Outcomes were any significant eQTL or sQTL. Logistic models included the standardized expression and splicing class scores, minor allele frequency and log TSS distance.

### Track-level predictions and cell classes

For the cellular context analyses we queried the Atlas RNA_SEQ scorer (371 tracks from 285 cell types and tissues) and the SPLICE_JUNCTIONS and SPLICE_SITE_USAGE scorers (367 tracks) for all 111,446 SNVs. We used the quantile layer, which ranks each predicted effect against a genome-wide variant background on a signed scale from −1 to 1. For each SNV and track we took the maximum absolute quantile over all genes (RNA-seq) or over all junctions and splice sites (splicing); 55.4% of SNVs had splicing predictions. Tracks were assigned to 12 cell classes by rule-based matching of biosample names, fixed before analysis. The classes were brain tissue; neurons (including neuroblastoma lines); glia (astrocytes and glioma lines); myeloid; lymphoid; mixed blood and immune (PBMC, blood, spleen); liver; metabolic and endocrine; vascular; muscle and heart; fibroblast and mesenchymal; and epithelial and other organs. For display, these were grouped into six super-classes (Fig. 5c).

### Entropy of predicted cellular labels

The score of an SNV for a class was the maximum absolute quantile over tracks in that class. This score was converted to a class-specific percentile across all SNVs, and the SNV’s predicted cellular label was the class with the highest percentile (ties broken by the raw maximum). Calibrating each class against its own distribution reduces differences in class scores arising from unequal numbers of tracks per class (5 to 137 tracks). For each locus we used uniform-prior credible-set variants weighted by PIP (weights summing to one within the locus) and computed *H* = −Σ_c_ π_c_ log πc, where π_c_ is the summed weight of variants with label *c*. Loci with an effective number of variants 1/Σ*w*^2^ < 2 were excluded. The null moved the whole set of credible-set variants circularly within the locus with their spacing preserved, replaced each position by the nearest SNV and kept the weights (1,000 replicates). The test was one-sided for lower entropy (greater concentration).

A cell-by-mechanism version used 24 categories. Each SNV was assigned to expression or splicing according to the larger calibrated percentile, with splicing percentiles computed among SNVs with splicing predictions. Sensitivity analyses gave each variant its top three classes with weight 1/3 each, used the uncalibrated top track, and restricted the analysis to variants with PIP ≥ 0.1. Secondary analyses measured pairwise agreement among the top 1% of variants in each locus (at least five) on the class and the exact biosample of the most affected track, and on the rank order of all tracks, defined as the Spearman correlation between the absolute-quantile profiles of the two variants across tracks (for each track, the maximum over genes). Pair means within loci were averaged over loci. The null translated the top set within its locus with its spacing preserved (1,000 replicates), and pairs were stratified by LD (*r*^2^ < 0.1 and *r*^2^ ≥ 0.5). Splicing versions used SNVs with splicing predictions and 367 tracks. Biosample agreement within LD strata and track rank order in high LD were computed post hoc (Supplementary Table S4). A further secondary analysis measured agreement between representative (highest-PIP) variants of different credible sets in the same locus, against a between-locus permutation null and, post hoc, a within-locus random-SNV null.

### Direction profiles

For each locus we selected the GWAS lead (minimum stage I *P*), credible-set members, the top 1% of SNVs by χ^2^ (at least five) and 80 random SNVs (fixed seed), 9,267 SNVs in total. We queried Atlas RNA_SEQ signed quantiles for all variant–gene pairs. Signs were oriented to the AD risk allele: *s* = sign(β) if the GWAS effect allele was the alternate allele and −sign(β) otherwise, and the risk-oriented effect was *s* × quantile. The target gene was the gene with the largest absolute quantile for the lead across tracks. The direction-profile concordance ρ_dir_ was the Spearman correlation between the 371-track risk-oriented profiles of a variant and of the lead for that gene; variants without a prediction for the gene were excluded. The locus statistic was the mean ρ_dir_ over compared variants, averaged over loci. The null replaced each compared variant with a random SNV of the same locus at a similar distance from the lead (distance ratio 0.5–2, otherwise the nearest distance; 1,000 replicates). Tests were one-sided.

The minority-sign fraction was computed for each SNV’s own most affected gene over tracks with |quantile| ≥ 0.5 (at least five such tracks). Secondary analyses compared the top 1% of variants outside credible sets and stratified variants by LD with the lead (*r*^2^ ≥ 0.5, 0.1–0.5, < 0.1). Sensitivity analyses used sign concordance (the fraction of same-sign tracks among tracks with |quantile| ≥ 0.5 for both variants), unoriented profiles, and genes strongly affected by both variants (|quantile| ≥ 0.9). For secondary credible sets (those not containing the lead), the representative was the highest-PIP variant. Its ρ_dir_ was expressed as a percentile of the ρ_dir_ values of distance-matched random SNVs of the same locus (median 12 per representative; range 3–56). This analysis was descriptive. Post hoc, the number above the 95th percentile was compared with its exact null distribution, in which each representative’s probability of exceeding the 95th percentile depends on its number of comparison SNVs (Poisson-binomial).

### Analysis protocols and sequence

Before examining the results of each subsequent stage, we documented its hypotheses, statistics, thresholds and sensitivity analyses in dated protocols, and amendments were recorded before results were inspected. These protocols were not registered externally. The exploratory stage specified the AVI–association test and the heterogeneity test as primary and the class-level analyses as descriptive. The confirmatory stage was specified after the exploratory results. It fixed one-sided directions for the expression and splicing classes (α = 0.025 each for the within-locus tests and the comparisons of highest-PIP with correlated variants). It also defined validity criteria for the new measurement (AUC ≥ 0.75 for missense and splice-region variants, and the *PILRA* control) and specified the genic-position and GTEx analyses. A relationship was considered not explained by genic position if, after adjustment, it remained in the pre-specified direction with one-sided *P* < 0.05; the GTEx tests used one-sided α = 0.05. The confirmatory stage passed six of seven primary tests: measurement validity, two within-locus tests, one comparison of highest-PIP with correlated variants and two GTEx tests. The splicing comparison of highest-PIP with correlated variants did not pass, and the splicing within-locus relationship did not meet the pre-specified genic-position criterion.

The cellular-context stage was specified before the track-level predictions were analysed. Its primary test was initially top-set class agreement; before results, an amendment made PIP-weighted entropy with class calibration the primary test and kept the other measures as secondary. The direction-profile analysis and the MPRA comparison were specified in separate protocols before their data were analysed; the MPRA methods are described in Supplementary Note 1. Neither primary hypothesis, greater concentration of predicted cellular labels or concordance of direction profiles with the lead, was supported.

## Data availability

GWAS summary statistics are available from the GWAS Catalog (GCST90027158). AlphaGenome Atlas scores [13] were obtained through the AlphaGenome API. GTEx v8 and Ensembl data were obtained through their public APIs. Microglial MPRA allelic-activity data were obtained from the repository accompanying Lee et al. [23] (github.com/RajLabMSSM/AD_MPRA). Per-variant AlphaGenome Atlas scores and the class scores derived from them are not redistributed with this article; they can be regenerated through the AlphaGenome API.

## Code availability

Analysis code is available at https://github.com/keras9496/ad-gwas-alphagenome-loci. Code that queries the AlphaGenome API and other external services is not included.

## Acknowledgements

We thank the Google DeepMind AlphaGenome team for making the AlphaGenome model and the precomputed AlphaGenome Atlas variant scores available to the research community. We thank the investigators of the European Alzheimer & Dementia Biobank (EADB) consortium and Bellenguez et al. for making their GWAS summary statistics publicly available, and Lee et al. for sharing their microglial MPRA data. The Genotype-Tissue Expression (GTEx) Project was supported by the Common Fund of the Office of the Director of the National Institutes of Health, and by NCI, NHGRI, NHLBI, NIDA, NIMH and NINDS. The data used for the analyses described in this manuscript were obtained from the GTEx Portal in September 2026.

## Funding

This research was supported by a grant of the Korea Health Technology R&D Project through the Korea Health Industry Development Institute (KHIDI), funded by the Ministry of Health & Welfare, Republic of Korea [grant number RS-2025-25455095].

## Author contributions

B.C. designed the study, performed the analyses and wrote the draft of the manuscript. J.J. supervised the study and reviewed the manuscript. J.J. and B.C. revised the manuscript.

## Conflicts of interest

The authors declare no conflicts of interest.

## Notes

### Competing Interest Statement

The authors have declared no competing interest.

### Author Declarations

The study used ONLY openly available human data that were publicly accessible, without application, screening or registration, before the initiation of the study. These data were originally located at: 1. Alzheimer's disease GWAS summary statistics (Bellenguez et al., 2022; stage I, GRCh38), GWAS Catalog accession GCST90027158: https://www.ebi.ac.uk/gwas/studies/GCST90027158 https://ftp.ebi.ac.uk/pub/databases/gwas/summary_statistics/GCST90027001-GCST90028000/GCST90027158/ 2. 1000 Genomes Project high-coverage (30x) GRCh38 genotypes, used as the LD reference panel (European individuals): https://www.internationalgenome.org/data-portal/data-collection/30x-grch38 http://ftp.1000genomes.ebi.ac.uk/vol1/ftp/data_collection/1000G_2504_high_coverage/ 3. GTEx v8 summary-level eQTL and sQTL associations, obtained through the public GTEx Portal and its API: https://gtexportal.org https://gtexportal.org/api/v2/ 4. Gene, exon and variant-consequence annotations from Ensembl (release 116) via the public Ensembl REST API and VEP: https://rest.ensembl.org 5. Microglial MPRA allelic-activity data from the public repository accompanying Lee et al.: https://github.com/RajLabMSSM/AD_MPRA AlphaGenome Atlas variant scores are computational predictions generated by the AlphaGenome model and do not contain data from human participants. No individual-level human data were accessed.

