## Supplementary Information for "Expression predictions inform molecular interpretation of Alzheimer’s disease risk loci"

**Contents:** Supplementary Note 1; Supplementary Tables S1–S4; Supplementary Figure S1. Reference numbers refer to the reference list of the main text.

### Supplementary Note 1. Exploratory comparison with microglial MPRA

**Rationale.** Within credible sets, the member with the highest AlphaGenome expression score might be more likely than the GWAS lead to show allelic regulatory activity. We tested this against allelic activity measured by MPRA of AD variants in the HMC3 microglial cell line and in iPSC-derived microglia (iMGL) [23].

**Methods.** Variants were linked by rsID. An emVar was defined, before analysis, as a variant in an active cis-regulatory sequence with allelic FDR < 0.05. The expression score was the confirmatory-stage raw-feature class score. The primary test (H1) used credible sets that contained the GWAS lead and at least two variants, in which the member with the highest expression score differed from the lead and both were tested (25 credible sets in each model). It compared the emVar status of the two variants with an exact one-sided McNemar test, with iMGL as the primary model and HMC3 as replication. Secondary tests were a conditional logistic regression of emVar status on the within-set standardized expression score across all tested credible-set members (H2), a comparison of the highest-expression member with a random tested member of the same set (H3; 10,000 draws), and the locus-wide AUC of the expression score with a locus-stratified permutation null (H4; 1,000 permutations).

**Results.** The highest-expression member was an emVar slightly more often than the lead in both models (iMGL 16% vs 12%; HMC3 12% vs 8%), but there were only five discordant pairs (3 vs 2; $P = 0.50$ in both; Supplementary Table S1). Across all tested variants, the expression score was associated with allelic activity in HMC3 cells (AUC 0.64, $P = 0.001$; within-set OR 1.50 per SD, $P = 0.004$) but not in iMGL (AUC 0.48; OR 0.99). One credible set was concordant in both models: at chr17:58.36 Mb near *MIR142*, the highest-expression member rs2526377 was an emVar in both HMC3 and iMGL, whereas the lead rs2680700 was an emVar in neither. The results were similar with alternative emVar thresholds, and the direction reversed in iMGL when the member was selected by the larger of the expression and splicing scores.

**Interpretation.** The exploratory MPRA comparison was inconclusive in iPSC-derived microglia, despite an association between expression scores and allelic activity in HMC3 cells; therefore, it does not provide consistent microglial validation of variant prioritization. These data neither support nor exclude the use of AlphaGenome expression scores to re-prioritize variants within credible sets. The discrepancy between HMC3 and iMGL may reflect the absence of microglia from AlphaGenome training data, or differences between the two MPRA models. MPRA measures episomal regulatory activity outside the endogenous chromatin context, and the MPRA library was selected by the authors' own fine-mapping.

**Supplementary Table S1 | Microglial MPRA comparison.** emVar, active cis-regulatory sequence with allelic FDR < 0.05. b/c, credible sets in which only the highest-expression member / only the lead is an emVar. One-sided *P* values. CS, credible set.

| Test | iMGL (primary) | HMC3 (replication) |
|---|---|---|
| H1: highest-expression member vs GWAS lead (25 CS) | emVar 0.16 vs 0.12; b/c 3/2; *P* = 0.50 | emVar 0.12 vs 0.08; b/c 3/2; *P* = 0.50 |
| H2: conditional logistic, OR per SD | 0.99 (0.79–1.23); *P* = 0.54; 37 CS, 568 variants | 1.50 (1.11–2.02); *P* = 0.004; 32 CS, 496 variants |
| H3: highest-expression vs random member (52 CS) | 0.096 vs 0.134; *P* = 0.86 | 0.173 vs 0.116; *P* = 0.12 |
| H4: locus-wide AUC (74 loci) | 0.48 (1,599 SNVs); *P* = 0.94 | 0.64 (1,597 SNVs); *P* = 0.001 |

**Supplementary Table S2 | Secondary credible sets exceeding the 95th percentile of matched comparisons in a post hoc screen.** Representative, highest-PIP variant of a credible set not containing the lead. Target gene, gene most affected by the lead across RNA-seq tracks. Percentile of $\rho_{dir}$ relative to distance-matched random SNVs of the same locus. *m*, number of distance-matched random SNVs; with fewer than 20, a percentile of 1.00 means that $\rho_{dir}$ exceeded all of them. Six of 35 secondary credible sets exceeded the 95th percentile, against 3.3 expected by chance (exact $P$ = 0.11). †, locus with suspected LD mismatch (Supplementary Table S3); the secondary credible set consisted of a single variant with PIP ≈ 1. Locus names follow the nearest established AD gene. These loci are hypotheses for subsequent validation; the profiles are model predictions, not measured effects.

| **Locus (hg38)** | **GWAS lead** | **Secondary representative** | **$r^2$ with lead** | **Target gene** | **$\rho_{dir}$** | **Percentile** | ***m*** |
|---|---|---|---|---|---|---|---|
| chr6:41.16 Mb (*TREM2* region) | rs143332484 (*TREM2* R62H) | rs4714447 | 0.00 | *TREM2* | 0.76 | 1.00 | 56 |
| chr14:92.47 Mb (*SLC24A4*) | rs12590654 | rs61977322 | 0.28 | *CHGA* | 0.57 | 1.00 | 15 |
| chr11:86.16 Mb (*PICALM*) | rs10792832 | rs3844143 | 0.39 | *RNU6-560P* | 0.53 | 1.00 | 8 |
| chr11:60.25 Mb (*MS4A*)† | rs1582763 | rs11230219 | 0.15 | *TCN1* | 0.41 | 1.00 | 10 |
| chr16:81.74 Mb (*PLCG2*) | rs12444183 | rs146121300 | 0.01 | ENSG00000261218 | 0.40 | 0.96 | 26 |
| chr16:30.01 Mb (*KAT8*)† | rs1140239 | rs12851 | 0.06 | *DOC2A* | 0.38 | 1.00 | 10 |

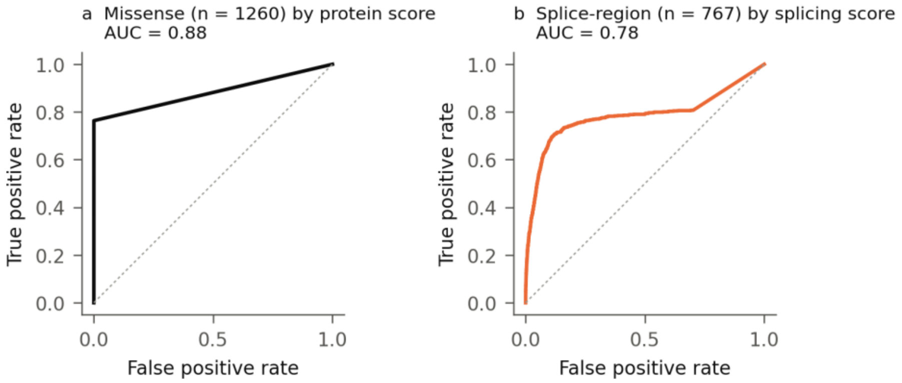


**Supplementary Figure S1 | Validity of raw-feature class scores. a,** ROC curve for VEP missense variants versus all other variants using the protein class score (*n* = 1,260 missense; AUC 0.88). Missense variants without an AlphaMissense value in the Atlas account for the plateau. **b,** ROC curve for VEP splice-region variants versus other genic variants using the splicing class score (n = 767 splice-region; AUC 0.78).

**Supplementary Table S3 | Analysed regions.** L001–L050, regions defined by genome-wide significant signals in the stage I data; R01–R25, additional regions reported for the same study in the GWAS Catalog (Methods). Each analysis window spans ±250 kb around the index variant (hg38). Index variant, stage I lead for L regions and Catalog-reported lead for R regions. Risk/other, risk allele (positive stage I β) and other allele; RAF, risk allele frequency; OR, odds ratio of the risk allele. Stage I *P*, *P* of the index variant in the stage I summary statistics; min. analysed *P*, smallest stage I *P* among analysed SNVs; Catalog *P*, *P* reported in the GWAS Catalog (stages I and II) for R regions. SNVs, analysed common SNVs (111,446 in total). Nearest gene, protein-coding gene whose transcription start site is nearest to the index variant (Ensembl release 116), which is not necessarily the gene implicated at the locus. CS, number of uniform-prior SuSiE credible sets. –, none.

| ID | Locus (hg38) | Index variant | Risk/other | RAF | OR | Stage I P | Min. analysed *P* | Catalog *P* | SNVs | Nearest gene | CS | Max PIP | Note |
|---|---|---|---|---|---|---|---|---|---|---|---|---|---|
| L001 | chr2:127.14 Mb | rs6733839 | T/C | 0.39 | 1.18 | $6.5 \times 10^{-90}$ | $6.5 \times 10^{-90}$ | – | 1,693 | *BIN1* | 3 | 1.00 | |
| L002 | chr11:86.16 Mb | rs10792832 | G/A | 0.64 | 1.11 | $6.3 \times 10^{-36}$ | $6.3 \times 10^{-36}$ | – | 1,414 | *EED* | 2 | 0.50 | |
| L003 | chr8:27.61 Mb | rs867230 | A/C | 0.60 | 1.11 | $1.5 \times 10^{-33}$ | $1.5 \times 10^{-33}$ | – | 1,689 | *SCARA3* | 2 | 0.83 | |
| L004 | chr1:207.52 Mb | rs6656401 | A/G | 0.19 | 1.13 | $2.8 \times 10^{-33}$ | $2.8 \times 10^{-33}$ | – | 1,228 | *CR1* | 1 | 0.52 | |
| L005 | chr19:1.05 Mb | rs12151021 | A/G | 0.34 | 1.11 | $4.1 \times 10^{-30}$ | $4.1 \times 10^{-30}$ | – | 2,019 | *ABCA7* | 2 | 1.00 | |
| L006 | chr11:60.25 Mb | rs1582763 | G/A | 0.63 | 1.09 | $1.7 \times 10^{-24}$ | $1.7 \times 10^{-24}$ | – | 1,515 | *MS4A4E* | 6 | 1.00 | LD mismatch suspected; excluded in sensitivity analysis |
| L007 | chr15:63.28 Mb | rs117618017 | T/C | 0.14 | 1.12 | $1.7 \times 10^{-21}$ | $1.7 \times 10^{-21}$ | – | 1,465 | *APH1B* | 2 | 0.98 | |
| L008 | chr6:41.16 Mb | rs143332484 | T/C | 0.01 | 1.40 | $6.0 \times 10^{-19}$ | $6.0 \times 10^{-19}$ | – | 1,664 | *TREM2* | 5 | 1.00 | |
| L009 | chr7:100.33 Mb | rs7384878 | T/C | 0.69 | 1.08 | $2.1 \times 10^{-18}$ | $2.1 \times 10^{-18}$ | – | 728 | *PILRB* | 1 | 0.49 | |
| L010 | chr2:233.12 Mb | rs10933431 | C/G | 0.77 | 1.09 | $1.0 \times 10^{-17}$ | $1.0 \times 10^{-17}$ | – | 1,763 | *INPP5D* | 2 | 0.95 | |
| L011 | chr17:63.48 Mb | rs4292 | T/C | 0.62 | 1.07 | $3.5 \times 10^{-16}$ | $3.5 \times 10^{-16}$ | – | 696 | *ACE* | 2 | 0.61 | |
| L012 | chr7:143.41 Mb | rs11769559 | T/C | 0.59 | 1.07 | $4.0 \times 10^{-16}$ | $4.0 \times 10^{-16}$ | – | 762 | *EPHA1* | 1 | 1.00 | |
| L013 | chr20:56.42 Mb | rs6014724 | A/G | 0.91 | 1.12 | $4.8 \times 10^{-16}$ | $4.8 \times 10^{-16}$ | – | 1,646 | *CASS4* | 1 | 0.21 | |
| L014 | chr14:92.47 Mb | rs12590654 | G/A | 0.67 | 1.07 | $2.1 \times 10^{-15}$ | $2.1 \times 10^{-15}$ | – | 1,847 | *RIN3* | 2 | 0.36 | |
| L015 | chr11:121.56 Mb | rs11218343 | T/C | 0.96 | 1.18 | $1.0 \times 10^{-14}$ | $1.0 \times 10^{-14}$ | – | 1,232 | *SORL1* | 3 | 1.00 | |
| L016 | chr10:80.49 Mb | rs6586028 | T/C | 0.80 | 1.08 | $1.3 \times 10^{-14}$ | $1.3 \times 10^{-14}$ | – | 1,703 | *TSPAN14* | 1 | 0.12 | |
| L017 | chr5:87.00 Mb | rs62375397 | T/C | 0.21 | 1.08 | $8.6 \times 10^{-14}$ | $8.6 \times 10^{-14}$ | – | 1,303 | *RASA1* | 1 | 0.12 | |
| L018 | chr4:11.02 Mb | rs6846529 | C/T | 0.28 | 1.07 | $1.3 \times 10^{-13}$ | $1.3 \times 10^{-13}$ | – | 1,928 | *CLNK* | 1 | 0.13 | |

| ID | Locus (hg38) | Index variant | Risk/other | RAF | OR | Stage I *P* | Min. analysed *P* | Catalog *P* | SNVs | Nearest gene | CS | Max PIP | Note |
|---|---|---|---|---|---|---|---|---|---|---|---|---|---|
| L019 | chr6:47.63 Mb | rs1385742 | A/T | 0.36 | 1.06 | $4.7 \times 10^{-13}$ | $4.7 \times 10^{-13}$ | – | 1,574 | *ADGRF4* | 1 | 0.07 | |
| L020 | chr17:49.22 Mb | rs616338 | T/C | 0.01 | 1.34 | $5.4 \times 10^{-13}$ | $5.4 \times 10^{-13}$ | – | 1,241 | *GNGT2* | 1 | 1.00 | |
| L021 | chr17:5.24 Mb | rs57402520 | A/G | 0.12 | 1.09 | $6.6 \times 10^{-13}$ | $6.6 \times 10^{-13}$ | – | 1,568 | *SCIMP* | 1 | 0.14 | |
| L022 | chr17:44.35 Mb | rs5848 | T/C | 0.29 | 1.07 | $1.8 \times 10^{-12}$ | $1.8 \times 10^{-12}$ | – | 980 | *GRN* | 1 | 1.00 | |
| L023 | chr16:81.74 Mb | rs12444183 | G/A | 0.61 | 1.06 | $2.2 \times 10^{-12}$ | $2.2 \times 10^{-12}$ | – | 2,479 | *PLCG2* | 4 | 0.72 | |
| L024 | chr10:11.68 Mb | rs7912495 | G/A | 0.46 | 1.06 | $2.9 \times 10^{-12}$ | $2.9 \times 10^{-12}$ | – | 1,695 | *USP6NL* | 1 | 0.97 | |
| L025 | chr8:144.10 Mb | rs34173062 | A/G | 0.08 | 1.12 | $2.9 \times 10^{-12}$ | $2.9 \times 10^{-12}$ | – | 767 | *MAF1* | 2 | 0.98 | |
| L026 | chr11:47.39 Mb | rs11500477 | A/T | 0.37 | 1.06 | $3.3 \times 10^{-12}$ | $3.3 \times 10^{-12}$ | – | 917 | *SLC39A13* | 1 | 0.29 | |
| L027 | chr5:180.20 Mb | rs113706587 | A/G | 0.11 | 1.10 | $3.4 \times 10^{-12}$ | $3.4 \times 10^{-12}$ | – | 1,858 | *RASGEF1C* | 1 | 0.33 | |
| L028 | chr16:30.01 Mb | rs1140239 | C/T | 0.62 | 1.06 | $4.6 \times 10^{-12}$ | $4.6 \times 10^{-12}$ | – | 513 | *C16orf92* | 6 | 1.00 | LD mismatch suspected; excluded in sensitivity analysis |
| L029 | chr15:58.76 Mb | rs602602 | T/A | 0.72 | 1.06 | $9.7 \times 10^{-12}$ | $9.7 \times 10^{-12}$ | – | 1,036 | *MINDY2* | 1 | 0.17 | |
| L030 | chr16:90.10 Mb | rs56407236 | A/G | 0.07 | 1.12 | $1.3 \times 10^{-11}$ | $1.3 \times 10^{-11}$ | – | 988 | *PRDM7* | 1 | 1.00 | |
| L031 | chr17:46.11 Mb | rs4510068 | G/T | 0.61 | 1.06 | $1.4 \times 10^{-10}$ | $1.4 \times 10^{-10}$ | – | 1,736 | *STH* | 1 | 0.09 | |
| L032 | chr14:105.73 Mb | rs28577986 | A/G | 0.26 | 1.07 | $3.4 \times 10^{-10}$ | $3.4 \times 10^{-10}$ | – | 373 | *TMEM121* | 1 | 0.36 | |
| L033 | chr4:0.99 Mb | rs3822030 | T/G | 0.57 | 1.05 | $5.0 \times 10^{-10}$ | $5.0 \times 10^{-10}$ | – | 1,774 | *SLC26A1* | 1 | 0.99 | |
| L034 | chr14:52.92 Mb | rs17125924 | G/A | 0.09 | 1.09 | $5.8 \times 10^{-10}$ | $5.8 \times 10^{-10}$ | – | 1,041 | *FERMT2* | 1 | 0.13 | |
| L035 | chr16:31.14 Mb | rs78924645 | G/A | 0.72 | 1.06 | $7.7 \times 10^{-10}$ | $7.7 \times 10^{-10}$ | – | 673 | *PRSS36* | 1 | 0.16 | |
| L036 | chr21:26.10 Mb | rs2154481 | T/C | 0.52 | 1.05 | $1.0 \times 10^{-9}$ | $1.0 \times 10^{-9}$ | – | 1,482 | *APP* | 2 | 0.24 | |
| L037 | chr8:94.99 Mb | rs4734295 | G/A | 0.46 | 1.05 | $2.0 \times 10^{-9}$ | $2.0 \times 10^{-9}$ | – | 1,541 | *TP53INP1* | 1 | 0.04 | |
| L038 | chr7:54.88 Mb | rs74504435 | A/G | 0.91 | 1.09 | $2.0 \times 10^{-9}$ | $2.0 \times 10^{-9}$ | – | 1,732 | *SEC61G* | 1 | 0.38 | |
| L039 | chr19:54.27 Mb | rs12984029 | A/G | 0.23 | 1.06 | $3.2 \times 10^{-9}$ | $3.2 \times 10^{-9}$ | – | 1,828 | *LILRB5* | 2 | 0.94 | |
| L040 | chr8:11.84 Mb | rs1065712 | C/G | 0.05 | 1.11 | $5.5 \times 10^{-9}$ | $5.5 \times 10^{-9}$ | – | 2,134 | *FDFT1* | 1 | 0.96 | |
| L041 | chr10:60.03 Mb | rs7068231 | G/T | 0.60 | 1.05 | $6.8 \times 10^{-9}$ | $6.8 \times 10^{-9}$ | – | 1,712 | *CCDC6* | 1 | 0.26 | |
| L042 | chr17:58.36 Mb | rs2680700 | G/T | 0.62 | 1.05 | $9.9 \times 10^{-9}$ | $9.9 \times 10^{-9}$ | – | 977 | *SUPT4H1* | 1 | 0.64 | |
| L043 | chr1:161.22 Mb | rs2070902 | C/T | 0.75 | 1.06 | $1.0 \times 10^{-8}$ | $1.0 \times 10^{-8}$ | – | 1,420 | *FCER1G* | – | – | |
| L044 | chr14:39.42 Mb | rs74745468 | A/G | 0.09 | 1.09 | $1.4 \times 10^{-8}$ | $1.4 \times 10^{-8}$ | – | 1,465 | *FBXO33* | 1 | 0.81 | |
| L045 | chr9:104.90 Mb | rs1800978 | G/C | 0.13 | 1.07 | $1.6 \times 10^{-8}$ | $1.6 \times 10^{-8}$ | – | 2,153 | *ABCA1* | 1 | 0.75 | |
| L046 | chr8:100.66 Mb | rs1693551 | C/T | 0.47 | 1.05 | $1.8 \times 10^{-8}$ | $1.8 \times 10^{-8}$ | – | 1,431 | *SNX31* | 1 | 0.26 | |
| L047 | chr15:49.90 Mb | rs183428733 | T/C | 0.01 | 1.27 | $1.8 \times 10^{-8}$ | $1.8 \times 10^{-8}$ | – | 1,418 | *SLC27A2* | 2 | 1.00 | |
| L048 | chr7:28.14 Mb | rs67250450 | T/C | 0.79 | 1.06 | $2.0 \times 10^{-8}$ | $2.0 \times 10^{-8}$ | – | 1,436 | *JAZF1* | 1 | 0.11 | |
| L049 | chr3:155.08 Mb | rs61762319 | G/A | 0.03 | 1.15 | $2.1 \times 10^{-8}$ | $2.1 \times 10^{-8}$ | – | 1,260 | *MME* | 1 | 0.60 | |
| L050 | chr6:114.36 Mb | rs976271 | A/G | 0.36 | 1.05 | $3.2 \times 10^{-8}$ | $3.2 \times 10^{-8}$ | – | 993 | *HS3ST5* | 1 | 0.16 | |

| ID | Locus (hg38) | Index variant | Risk/other | RAF | OR | Stage I *P* | Min. analysed *P* | Catalog *P* | SNVs | Nearest gene | CS | Max PIP | Note |
|---|---|---|---|---|---|---|---|---|---|---|---|---|---|
| R01 | chr2:105.75 Mb | rs143080277 | C/T | 0.005 | 1.48 | $7.9 \times 10^{-11}$ | $7.0 \times 10^{-6}$ | $2.0 \times 10^{-13}$ | 2,267 | *NCK2* | – | – | Index variant not analysed (MAF < 1%) |
| R02 | chr7:12.23 Mb | rs13237518 | C/A | 0.59 | 1.04 | $5.1 \times 10^{-7}$ | $3.9 \times 10^{-7}$ | $5.0 \times 10^{-11}$ | 2,699 | *TMEM106B* | 1 | 0.02 | |
| R03 | chr10:96.27 Mb | rs6584063 | A/G | 0.96 | 1.12 | $1.8 \times 10^{-7}$ | $1.1 \times 10^{-7}$ | $7.0 \times 10^{-11}$ | 1,359 | *BLNK* | 1 | 0.06 | |
| R04 | chr17:1.73 Mb | rs35048651 | T/TGAG | 0.21 | 1.05 | $4.8 \times 10^{-7}$ | $2.4 \times 10^{-6}$ | $8.0 \times 10^{-11}$ | 1,733 | *WDR81* | 1 | 0.05 | Index variant not analysed (indel) |
| R05 | chr7:7.82 Mb | rs6943429 | T/C | 0.42 | 1.04 | $3.0 \times 10^{-7}$ | $2.3 \times 10^{-7}$ | $1.0 \times 10^{-10}$ | 2,106 | *RPA3* | 1 | 0.23 | |
| R06 | chr7:37.84 Mb | rs6966331 | C/T | 0.65 | 1.04 | $4.8 \times 10^{-6}$ | $2.2 \times 10^{-7}$ | $5.0 \times 10^{-10}$ | 1,751 | *NME8* | – | – | |
| R07 | chr16:70.66 Mb | rs4985556 | A/C | 0.11 | 1.06 | $5.6 \times 10^{-6}$ | $3.4 \times 10^{-6}$ | $6.0 \times 10^{-10}$ | 953 | *MTSS2* | – | – | |
| R08 | chr17:18.16 Mb | rs2242595 | G/A | 0.89 | 1.06 | $4.6 \times 10^{-6}$ | $3.7 \times 10^{-6}$ | $1.0 \times 10^{-9}$ | 992 | *ALKBH5* | – | – | |
| R09 | chr4:40.20 Mb | rs2245466 | G/C | 0.34 | 1.05 | $3.1 \times 10^{-7}$ | $3.1 \times 10^{-7}$ | $1.0 \times 10^{-9}$ | 1,707 | *RHOH* | 1 | 0.99 | |
| R10 | chr12:113.28 Mb | rs6489896 | C/T | 0.08 | 1.08 | $2.5 \times 10^{-6}$ | $2.5 \times 10^{-6}$ | $2.0 \times 10^{-9}$ | 1,243 | *DRC10* | 1 | 0.01 | |
| R11 | chr5:14.72 Mb | rs112403360 | A/T | 0.07 | 1.08 | $3.4 \times 10^{-6}$ | $2.8 \times 10^{-7}$ | $2.0 \times 10^{-9}$ | 1,306 | *OTULIN* | 1 | 0.14 | |
| R12 | chr20:63.74 Mb | rs6742 | C/T | 0.78 | 1.05 | $2.5 \times 10^{-6}$ | $2.5 \times 10^{-6}$ | $3.0 \times 10^{-9}$ | 1,869 | *SLC2A4RG* | – | – | |
| R13 | chr2:37.30 Mb | rs17020490 | C/T | 0.14 | 1.06 | $3.3 \times 10^{-6}$ | $5.0 \times 10^{-7}$ | $3.0 \times 10^{-9}$ | 1,873 | *PRKD3* | – | – | |
| R14 | chr10:122.41 Mb | rs7908662 | A/G | 0.53 | 1.04 | $3.3 \times 10^{-6}$ | $2.6 \times 10^{-6}$ | $3.0 \times 10^{-9}$ | 1,696 | *PLEKHA1* | 1 | 0.05 | |
| R15 | chr15:78.94 Mb | rs12592898 | G/A | 0.87 | 1.06 | $1.3 \times 10^{-6}$ | $1.3 \times 10^{-6}$ | $4.0 \times 10^{-9}$ | 1,636 | *CTSH* | 1 | 0.40 | |
| R16 | chr19:49.95 Mb | rs9304690 | T/C | 0.24 | 1.05 | $6.4 \times 10^{-6}$ | $5.7 \times 10^{-6}$ | $5.0 \times 10^{-9}$ | 1,180 | *SIGLEC11* | 1 | 0.22 | |
| R17 | chr1:109.35 Mb | rs141749679 | C/T | 0.004 | 1.37 | $8.0 \times 10^{-6}$ | $8.3 \times 10^{-6}$ | $8.0 \times 10^{-9}$ | 1,148 | *MYBPHL* | – | – | Index variant not analysed (MAF < 1%) |
| R18 | chr5:151.05 Mb | rs871269 | C/T | 0.67 | 1.04 | $3.4 \times 10^{-6}$ | $3.4 \times 10^{-6}$ | $9.0 \times 10^{-9}$ | 1,818 | *GPX3* | – | – | |
| R19 | chr16:86.42 Mb | rs16941239 | A/T | 0.03 | 1.13 | $2.9 \times 10^{-6}$ | $2.9 \times 10^{-6}$ | $1.0 \times 10^{-8}$ | 2,619 | *FOXF1* | – | – | |
| R20 | chr2:202.88 Mb | rs139643391 | TC/T | 0.87 | 1.07 | $2.6 \times 10^{-7}$ | $2.8 \times 10^{-7}$ | $1.0 \times 10^{-8}$ | 825 | *ICA1L* | 1 | 0.01 | Index variant not analysed (indel) |
| R21 | chr20:0.41 Mb | rs1358782 | G/A | 0.75 | 1.05 | $1.9 \times 10^{-6}$ | $6.7 \times 10^{-7}$ | $2.0 \times 10^{-8}$ | 1,850 | *RBCK1* | – | – | |
| R22 | chr16:79.57 Mb | rs450674 | T/C | 0.63 | 1.05 | $1.1 \times 10^{-7}$ | $1.1 \times 10^{-7}$ | $3.0 \times 10^{-8}$ | 2,283 | *MAF* | 1 | 0.28 | |
| R23 | chr2:9.56 Mb | rs72777026 | G/A | 0.14 | 1.06 | $1.9 \times 10^{-6}$ | $7.0 \times 10^{-8}$ | $3.0 \times 10^{-8}$ | 1,304 | *ADAM17* | 1 | 0.11 | |
| R24 | chr4:139.59 Mb | rs535347112 | T/C | 0.001 | 2.15 | $5.8 \times 10^{-8}$ | $6.3 \times 10^{-5}$ | $4.0 \times 10^{-8}$ | 1,335 | *SETD7* | – | – | Index variant not analysed (MAF < 1%) |
| R25 | chr17:7.56 Mb | rs58840546 | TCAA/T | 0.94 | 1.09 | $2.1 \times 10^{-7}$ | $7.3 \times 10^{-7}$ | $5.0 \times 10^{-8}$ | 1,404 | *SENP3* | 1 | 1.00 | Index variant not analysed (indel) |

**Supplementary Table S4 | Pairwise agreement among top associated variants on the most affected track.** Pairs were formed among the top 1% of SNVs by $\chi^2$ in each locus (at least five). Class and biosample, fraction of pairs whose most affected track belongs to the same cell class (12 classes) or the same biosample; track rank order, Spearman correlation between the absolute-quantile profiles of the two variants across all tracks (for each track, the maximum over genes). Pair means within loci were averaged over loci. Null mean, mean over 1,000 sets obtained by translating the top set within its locus with its spacing preserved. One-sided *P* for greater agreement than the null. Low LD, $r^2 < 0.1$; high LD, $r^2 \geq 0.5$ (1000 Genomes European); pairs with $0.1 \leq r^2 < 0.5$ are in neither stratum. Splicing analyses used SNVs with splicing predictions (55% of SNVs) and 367 tracks. Secondary, pre-specified secondary analysis; post hoc, computed after the original analysis with the same procedure.

| Measure | LD stratum | Observed | Null mean | One-sided *P* | Loci | Pairs | Analysis |
|---|---|---|---|---|---|---|---|
| **RNA-seq predictions (371 tracks)** | | | | | | | |
| Cell class of most affected track | All pairs | 0.201 | 0.224 | 0.97 | 75 | 8,554 | Secondary |
| | Low ($r^2 < 0.1$) | 0.163 | 0.190 | 0.86 | 37 | 1,908 | Secondary |
| | High ($r^2 \geq 0.5$) | 0.211 | 0.243 | 1.00 | 75 | 5,065 | Secondary |
| Exact biosample of most affected track | All pairs | 0.024 | 0.055 | 1.00 | 75 | 8,554 | Secondary |
| | Low ($r^2 < 0.1$) | 0.014 | 0.024 | 0.95 | 37 | 1,908 | Post hoc |
| | High ($r^2 \geq 0.5$) | 0.025 | 0.076 | 1.00 | 75 | 5,065 | Post hoc |
| Track rank order (mean Spearman ρ) | All pairs | 0.132 | 0.155 | 1.00 | 75 | 8,554 | Secondary |
| | Low ($r^2 < 0.1$) | 0.116 | 0.112 | 0.36 | 37 | 1,908 | Secondary |
| | High ($r^2 \geq 0.5$) | 0.139 | 0.179 | 1.00 | 75 | 5,065 | Post hoc |
| **Splicing predictions (367 tracks)** | | | | | | | |
| Cell class of most affected track | All pairs | 0.287 | 0.359 | 1.00 | 73 | 2,994 | Secondary |
| | Low ($r^2 < 0.1$) | 0.245 | 0.279 | 0.76 | 33 | 734 | Secondary |
| | High ($r^2 \geq 0.5$) | 0.303 | 0.397 | 1.00 | 71 | 1,766 | Secondary |
| Exact biosample of most affected track | All pairs | 0.113 | 0.205 | 1.00 | 73 | 2,994 | Secondary |
| | Low ($r^2 < 0.1$) | 0.042 | 0.128 | 1.00 | 33 | 734 | Post hoc |
| | High ($r^2 \geq 0.5$) | 0.115 | 0.245 | 1.00 | 71 | 1,766 | Post hoc |
| Track rank order (mean Spearman ρ) | All pairs | 0.251 | 0.320 | 1.00 | 73 | 2,994 | Secondary |
| | Low ($r^2 < 0.1$) | 0.215 | 0.199 | 0.33 | 33 | 734 | Secondary |
| | High ($r^2 \geq 0.5$) | 0.270 | 0.371 | 1.00 | 71 | 1,766 | Post hoc |

94
